# Peptide receptor radionuclide therapy targeting the cholecystokinin-2 receptor: Preclinical and first clinical experience in small cell lung cancer

**DOI:** 10.1101/2025.07.30.25330817

**Authors:** Taraneh Sadat Zavvar, Giulia Santo, Leonhard Gruber, Ariane Kronthaler, Judith Hagenbuchner, Ira Skvortsova, Inken Piro, Katja Steiger, Vladan Martinovic, Daniela Minasch, Judith Löffler-Ragg, Gianpaolo di Santo, Irene J. Virgolini, Elisabeth von Guggenberg

## Abstract

**Rationale:** Peptide receptor radionuclide therapy (PRRT) is an established treatment for neuroendocrine tumors (NETs), enabling targeted radiation delivery via radiolabeled peptides. Small cell lung cancer (SCLC) remains a major therapeutic challenge due to its aggressive nature and poor prognosis. Despite advances, relapse rates are high and effective therapies are limited. We previously demonstrated the diagnostic potential of the cholecystokinin-2 receptor-targeting minigastrin analog [^68^Ga]Ga-DOTA-MGS5 in PET/CT imaging in different NETs. Building on this, we developed and evaluated [^177^Lu]Lu-DOTA-MGS5 as a therapeutic PRRT agent.

**Methods:** Preclinical studies investigating the cellular internalization and intracellular distribution over time in A431 cells were performed using the fluorescent tracer ATTO-488-MGS5. Short-and long-term cytotoxic effects of [^177^Lu]Lu-DOTA-MGS5 were evaluated on the same cell line using trypan blue exclusion and clonogenic survival assays. CCK2R expression was assessed by immunohistochemistry in 42 SCLC tissue specimens. In addition, the first PRRT with [^177^Lu]Lu-DOTA-MGS5 was conducted in a patient with extensive disease SCLC after confirming CCK2R-positive uptake in [^68^Ga]Ga-DOTA-MGS5 PET/CT.

**Results:** Rapid binding and internalization into A431-CCK2R cells, with progressive accumulation in intracellular compartments, was observed for ATTO-488-MGS5. Short-term irradiation effects of [^177^Lu]Lu-DOTA-MGS5 were comparable for 4 h and 24 h incubation and lay between the effects obtained with 2 and 4 Gy of EBRT. Clonogenic survival of A431-CCK2R cells incubated with the increasing radioactivity of [^177^Lu]Lu-DOTA-MGS5 decreased in a dose-dependent manner.

Immunohistochemistry on SCLC specimens confirmed moderate to high CCK2R expression in 16 of 42 SCLC samples. In the first patient with SCLC treated with four cycles of [^177^Lu]Lu-DOTA-MGS5 with a total activity of 17.2 GBq, no signs of toxicity were noticed.

**Conclusions:** The preclinical and clinical results confirm the feasibility of [^177^Lu]Lu-DOTA-MGS5 PRRT in patients with SCLC and support further clinical studies investigating the therapeutic value and clinical applicability of this new CCK2R-targeted theranostic approach in larger patient cohorts.

## Introduction

Peptide receptor radionuclide therapy (PRRT) has made a significant progress as a highly effective treatment modality of various cancers, in particular neuroendocrine tumors (NETs). PRRT employs radiolabeled peptides that bind to specific receptors, enabling targeted delivery of cytotoxic radiation directly to cancer cells, thus minimizing damage to surrounding healthy tissues (1).

With the approval of [^177^Lu]Lu-DOTA-TATE (Lutathera®) targeting somatostatin receptors (SSTR) by the European Medicines Agency (EMA) in 2017 and the FDA in 2018, following the positive outcomes of the NETTER-1 phase III trial, a broader clinical application of PRRT has become possible. The study demonstrated significantly improved progression-free survival in patients with advanced midgut NETs treated with [^177^Lu]Lu-DOTA-TATE (2). In various studies significant improvements in patient outcomes with PRRT, in terms of overall response rates and quality of life were shown (3–5).

Lung neuroendocrine neoplasms (NENs) represent a heterogeneous group of tumors characterized by neuroendocrine morphology and immunophenotype. According to the 2022 WHO classification, they are divided into well-differentiated NETs—including typical carcinoid (low-grade) and atypical carcinoid (intermediate-grade) as well as NETs with elevated mitotic counts and/or proliferation index—and poorly differentiated neuroendocrine carcinomas (NECs), which comprise small cell (lung) carcinoma and large cell neuroendocrine carcinoma (6).

Notably, in contrast to gastroenteropancreatic neuroendocrine tumors (GEP-NETs), SSTR expression in pulmonary NETs shows greater heterogeneity. Recent studies report that only 24–44% of patients with metastatic pulmonary NET demonstrate uniformly positive SSTR expression on positron emission tomography (PET) imaging (7–9). The expression of SSTR was analyzed by immunohistochemistry in SCLC tissue samples, revealing that 38% of the specimens were positive for SSTR subtype 2 (10). Consequently, there is a clear need to investigate alternative radiopharmaceuticals beyond those targeting SSTRs, in order to provide effective diagnostic and therapeutic options for patients lacking sufficient SSTR expression.

Small cell lung cancer (SCLC) accounting for 13% to 15% of all lung cancer cases globally represents a significant clinical challenge. Extensive disease is associated with a particularly poor prognosis, with a five-year survival rate commonly reported to be less than 10% (11–13). The treatment strategies for SCLC have evolved, with the current standard of care primarily involving a combination of platinum-based chemotherapy - either cisplatin or carboplatin - paired with etoposide. This regimen is recommended by several guidelines, for both limited disease (LD) and extensive disease (ED) SCLC (13–15). Recent updates have incorporated the use of immune checkpoint inhibitors such as atezolizumab or durvalumab in conjunction with standard chemotherapy, particularly for patients with ED-SCLC, reflecting a shift towards more comprehensive therapeutic approaches (15, 16). The management of SCLC has been shown to benefit from additional interventions such as prophylactic cranial irradiation (PCI), especially in responding patients, as it can significantly reduce the risk of symptomatic brain metastases (17).

Despite advancements in cancer treatment, there remains a significant need to improve therapeutic options and explore novel treatment modalities for patients with SCLC, as current therapies often provide limited efficacy and are associated with high relapse rates. Furthermore, emerging strategies focusing on personalized approaches, may pave the way for more effective and individualized treatment paradigms, potentially improving prognosis in this challenging disease (18, 19). Therefore, the exploration of novel agents, including radiopharmaceuticals and combination therapies, is essential for future research and clinical application in the management of SCLC (20, 21).

Previously, we have demonstrated the ability of the new radiolabeled minigastrin analogue [^68^Ga]Ga-DOTA-MGS5 targeting the cholecystokinin-2 receptor (CCK2R) to accurately detect malignant lesions by means of PET/CT imaging, supporting its potential as a valuable diagnostic tool in patients with advanced medullary thyroid carcinoma (MTC) as well as SCLC (22, 23). Within a phase I/IIA clinical trial the safety and dosimetry, as well as the diagnostic performance of [^68^Ga]Ga-DOTA-MGS5 was evidenced in patients with advanced MTC and a patient with bronchopulmonary NET (24). In addition, we have reported the successful radiopharmaceutical development and preclinical evaluation of [^177^Lu]Lu-DOTA-MGS5, establishing a solid foundation for its further advancement as a therapeutic agent (25).

Within this study, we report on the CCK2R expression in tumor samples from patients with SCLC evaluated by immunohistochemistry. We further investigated the receptor interaction at the cellular level using the peptide analog MGS5 conjugated to a fluorescence dye. The therapeutic effects of [^177^Lu]Lu-DOTA-MGS5 were also assessed by comparing CCK2R expressing cells incubated with [^177^Lu]Lu-DOTA-MGS5 with the same cell lines treated with external beam radiotherapy (EBRT). In addition, we report on the first application of [^177^Lu]Lu-DOTA-MGS5 in a patient with ED-SCLC. The results provide the preliminary basis for future clinical trials in this patient group.

## Materials and Methods

### Cell cultures

The A431 human epidermoid carcinoma cell line stably transfected with the plasmid pCR3.1 containing the complete human CCK2R coding sequence and the same cell line transfected with the empty vector alone were originally provided by Dr. Luigi Aloj (26). A431 cells were grown in Dulbecco’s modified Eagle’s medium (DMEM) supplemented with 10% (v/v) foetal bovine serum and 5 mL of a 100x penicillin-streptomycin-glutamine mixture. Cells were cultured in a humidified atmosphere containing 5% CO_2_ at 37°C. Cells were passaged three times per week with 10x 2.5% trypsin-EDTA solution at a 1:2-1:3 ratio. All media and supplements were purchased from Sigma-Aldrich (Darmstadt, Germany) or Invitrogen Corporation (Lofer, Austria).

### Fluorescence microscopy

ATTO-488 NHS-ester (ATTO-TEC, Siegen, Germany) was conjugated to the N-terminus of the MGS5 peptide sequence via amide bound. The reaction was carried out in acetonitrile under basic conditions (10 µL of DIPEA, pH∼9) for 1 h. After coupling, ATTO-488-MGS5 was purified by preparative HPLC, lyophilized and stored at-20°C. Conjugation was confirmed by mass analysis using Q Exactive HF mass spectrometer (Thermo Fisher Scientific, Bremen, Germany). The structural formula of ATTO-488-MGS5 (MW: 1635 g/mol) is shown in Figure 1A.

**Figure 1.**
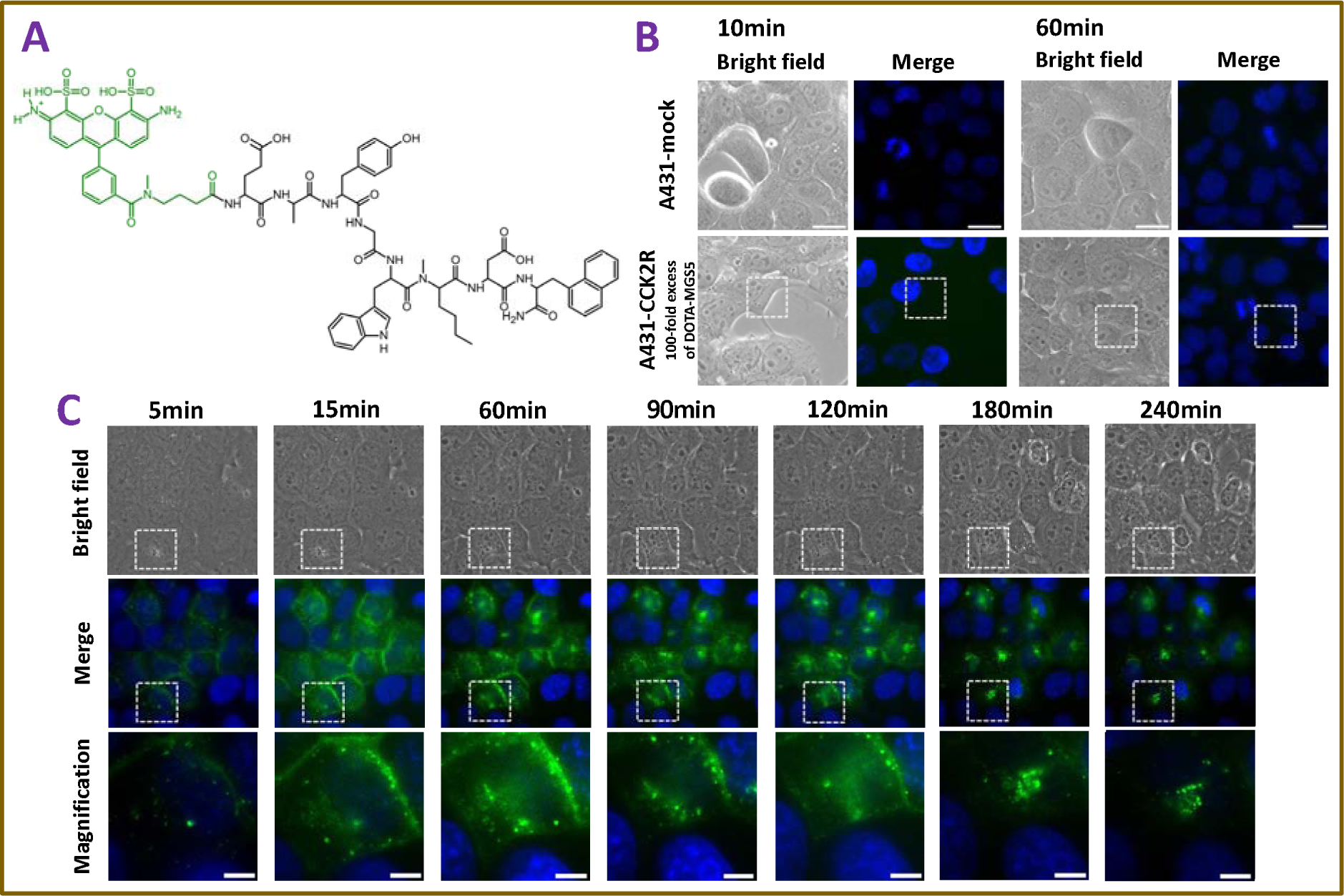
Fluorescence imaging of A431-CCK2R and A431-mock cells. (A) Structural formula of ATTO-488-MGS5 (B) Uptake of ATTO-488-MGS5 in A431-mock cells after 10 min and 60 min of incubation (upper row). The lower row shows uptake of A431-CCK2R cells with pre-incubation of DOTA-MGS5 (10 µM) as a blocking agent before ATTO-488-MGS5 exposure. (C) Time-dependent uptake of ATTO-488-MGS5 in A431-CCK2R from 5 min to 240 min. Cell nuclei are stained with Hoechst (blue), and ATTO488-MGS5 (100 nM) is shown in green. Image size: 90 × 90 µm, magnification: 25 × 25 µm, scale bar: 5 µm.

The cell uptake of the fluorescent labeled tracer was analyzed on A431-CCK2R and A431-mock cells. Cells were seeded on labteks or glass plates coated with 0.1 mg/mL collagen (Corning, New York, USA) at a density of 30.000 or 500.000 cells, 36 to 48 h prior to assay. In order to differentiate the nucleus, nuclei were stained using 1 µg/mL Hoechst 33342 (Merck, Vienna, Austria) 30 min before imaging. Then cells were incubated with 100 nM of ATTO-488-MGS5 and cellular microscopy was performed using an Axiovert 200M microscope (Zeiss, Oberkochen, Germany), using a setting of λ_exc_ = 405 nm for Hoechst nuclear staining, and λ_exc_ = 488 nm for the fluorescent ligand. Image analyses were done with Axiovision Software (Zeiss, Oberkochen, Germany). Receptor-specific uptake in A431-CCK2R cells was confirmed by additional blocking studies with 100-fold excess of DOTA-MGS5 (10 µM) and using A431-mock cells without receptor expression as negative control.

### Short-Term cell cytotoxicity assay

To investigate the cytotoxic effect of [^177^Lu]Lu-DOTA-MGS5 on A431-CCK2R cells in comparison with EBRT, cells were counted using a TC20 automated cell counter (Bio-Rad Laboratories, Hercules, CA, USA), seeded at a density of 1 × 10⁵ cells per well in 6-well plates and incubated at 37°C with 5% CO₂. After 24 h, cells were irradiated with varying doses 2, 4 and 8 Gy using a linear accelerator (Elekta Precise, Elekta Oncology Systems, UK) or by incubation with [^177^Lu]Lu-DOTA-MGS5 (∼250 kBq, 2nM/well) for 4 and 24 h followed by replacement with fresh medium.

At the time point of 72 h after treatment, cell viability was assessed using a Beckman Coulter Vi-CELL AS cell viability analyzer (Beckman Coulter, Fullerton, CA, USA). The supernatant from each well was collected in 15 mL Falcon tubes. Cells were then washed twice with 1 mL of serum-free medium, and the wash fractions were combined with the respective supernatants. Trypsin (300 µL) was added to each well, and plates were incubated until detachment. Following detachment, 1 mL of medium was added to each well, and cells were rinsed and transferred to their respective Falcon tubes. This washing step was repeated twice reaching a total volume of 3 mL.

Cells were pelleted by centrifugation in a benchtop centrifuge (Megafuge™ 8, Thermo Fisher Scientific, Waltham, MA, USA) at 2000 rpm for 8 min at 4°C, and the supernatant was discarded. Pellets were resuspended in 1 mL of cold serum-free medium. Samples were loaded into the Vi-CELL AS analyzer according to the manufacturer’s instructions. The number of viable cells in each treatment group was normalized to the mean number of viable cells in the non-irradiated control group and expressed as a percentage of the control. The average diameter and circularity of cells in each group were analyzed using ordinary one-way ANOVA.

### Clonogenic Survival Assay

To investigate the clonogenic survival after incubation with [^177^Lu]Lu-DOTA-MGS5, cells were counted in the automated cell counter and serial dilutions were performed to achieve a final concentration of 1,000 cells/mL (1 cell/µL). Based on the treatment with increasing activities of [^177^Lu]Lu-DOTA-MGS5 (250 kBq-2 MBq, ∼100 nM in each well), cell numbers seeded in 6-well plates were adapted and incubated at 37°C with 5% CO₂. After 24 h of incubation to allow for cell attachment, cells were treated with the increasing activities of [^177^Lu]Lu-DOTA-MGS5 for 24 h at 37°C. After this treatment, the medium was aspirated, the cells were gently washed twice with phosphate-buffered saline (PBS) and then incubated at 37°C for 12 days to allow colony formation. At the end of the incubation period, the culture medium was removed, and the wells were gently rinsed twice with PBS.

Colonies were fixed by adding 1 mL of a 3:1 methanol:glacial acetic acid solution to each well and incubated for 20 min at room temperature in a chemical hood. The solution was removed and the fixed cells gently rinsed twice with 1 mL of PBS.

Staining was performed using 1 mL of Giemsa stain (modified solution; Sigma-Aldrich, CAS No. 32884) diluted 3.5:10 in Milli-Q water. Plates were incubated with the staining solution for 2 h at room temperature. After staining, wells were rinsed twice with Milli-Q water, and plates were inverted on absorbent paper to air dry.

Colonies were counted manually, with each visible purple-stained cluster considered as one colony. The number of colonies in each treatment group was normalized to the plating efficiency of a control group not subjected to incubation with [^177^Lu]Lu-DOTA-MGS5, and expressed as percentage of clonogenic survival. Each data point represents the mean ± standard deviation (SD) of four independent experiments, each performed in triplicates.

### Immunohistochemistry

The expression of CCK2R in SCLC was assessed by immunohistochemistry on tissue specimens available at the tissue archive of the Institute of Pathology of the Technical University of Munich, Germany. The Medical Ethics Committee of the Technical University of Munich gave ethical approval for this work (2024-130-S-DFG-SB). A total of 42 tissue specimens, each from a distinct patient (25 men and 17 women) with histologically confirmed SCLC, were analyzed. The samples comprised tissue samples taken from 41 primary tumors and one lymph node metastasis. Slides were deparaffinized and rehydrated using decreasing concentration of alcohol, followed by heat induced epitope retrieval at 100 °C for 40 min in Bond™ H1 solution. Endogenous peroxidase activity was blocked, and sections were incubated with a mouse monoclonal CCK2R (E-3) antibody raised against amino acids 1-85 mapping at the N-terminus of CCK2R of human origin (sc-166690, Santa Cruz Biotechnology, Dallas, TX, USA) at a dilution of 1:75 for 15 min at room temperature.

Bond Polymer Refine Detection Kit (DS9800, Leica Biosystems, Wetzlar, Germany) without the post-primary reagent was used for antibody detection, followed by DAB (3,3’-diaminobenzidine, BS04-500, Medac Diagnostica, Wedel, Germany) staining using brown chromogen (DS9800, Bond Polymer Refine Detection, Leica Biosystems, Wetzlar, Germany), hematoxylin counterstaining, dehydration, clearing, and mounting using Pertex mounting medium (00801, Histolab, Goeteborg, Sweden). Slides were digitally scanned with a resolution of 0.252 µm per pixel (equivalent to a 40x objective) using a Leica Aperio Imager AT2 (Leica Biosystems, Wetzlar, Germany). Images were analyzed using AperioImageScope software. The same procedure was applied to the tissue specimen of the patient with ED-SCLC treated with [^177^Lu]Lu-DOTA-MGS5. Antibody specificity was evaluated using specimens from human stomach tissue, known to physiologically express CCK2R, and normal lung tissue lacking CCK2R expression (Supplementary Figure 1).

All samples were evaluated for CCK2R immunoreactivity in terms of a positive signal (brown DAB precipitate, Figure 3). Staining intensity (I, score 0–3) and frequency (P, %) was assessed for each tumor sample. A final immunoreactivity score was defined as IRS = I × (P/100) – with IRS being in a final range between 0 and 3.

### Clinical case presentation

A patient diagnosed with SCLC (initial stage: T4 N3 M1b with primary tumor of the left central lung and possible metastasis to the left adrenal gland) completed first-line systemic therapy with carboplatin/etoposide/atezolizumab (schedule day 1–day 3, every three weeks) followed by maintenance therapy with atezolizumab and prophylactic cranial irradiation (fractions of 2 Gy each, reaching a total dose of 30 Gy; cognitive preservation with memantine). Subsequently, the entire primary tumor region, including the selectively involved lymph nodes on the left central side, was irradiated (fractions of 2 Gy, resulting in a total dose of 60 Gy). After completing EBRT approximately six months after diagnosis, the disease remained stable for the following four months. Ten months after diagnosis, a new metastasis to the right adrenal gland occurred, and re-induction therapy with carboplatin/etoposide/atezolizumab was administered (a total of four cycles, schedule day 1–day 3, every three weeks). In the follow-up CT scan, progressive disease was identified again and third-line therapy with cyclophosphamide/doxorubicin/vincristine was initiated (a total of four cycles, schedule day 1–day 2, every three weeks). Further progression was seen at the next follow-up CT scan and the therapy was switched to fourth-line treatment with weekly topotecan (day 1, day 8, day 15).

After 2 cycles, the disease again progressed. This time, there was progression of the primary tumor, liver metastases, and new lymph node metastases. Thus, alternative image-guided treatment strategies were evaluated. PET/CT imaging with [^68^Ga]Ga-DOTA-MGS5 targeting the CCK2R, performed seventeen months after initial diagnosis, showed intense uptake in tumor lesions, while only faint somatostatin receptor-related uptake was observed in PET imaging with [^68^Ga]Ga-DOTA-TOC (23). Thus, the patient was considered eligible for PRRT with [^177^Lu]Lu-DOTA-MGS5. PRRT was performed under the responsibility of the treating physician and after tumor board approval. According to the Austrian Medicines Act (Arzneimittelgesetz §8) the named patient use (“Heilversuch”) was not subject to authorisation by or notification to the authorities. Written informed consent was obtained from the patient. All procedures were in accordance with the principles of the 1964 Declaration of Helsinki and its subsequent amendments. The retrospective analysis of the data of the patient was exempted from consultation with the Ethics Committee by the Medical University of Innsbruck.

### [^177^Lu]Lu-DOTA-MGS5 PRRT

#### Automated radiolabeling process

A dose of 1.500 MBq for dosimetric evaluation and four therapeutic individual doses of 4000 MBq [^177^Lu]Lu-DOTA-MGS5 were prepared using an automated synthesis module (Modular-Lab PharmTracer®, Eckert & Ziegler Eurotrope GmbH, Berlin, Germany) following a previously published method (25). Each batch was subjected to a full quality control evaluation before release for patient administration. The specifications for the final product which were established based on experience with in-house production of different radiotherapeutics and Ph. Eur. monographs available for other radiopharmaceuticals (27), are shown in Supplementary Table 1.

#### Treatment Regimen

Adequate hydration was ensured through intravenous infusion of 1000 mL of 0.9% saline solution at a rate of 300 mL/h, starting 30 min before and continuing for 2 h after treatment administration. The patient received prophylactic antiemetic therapy with 4 mg ondansetron, as well as 4 mg dexamethasone and 40 mg pantoprazole, given 20 min before the treatment. Prophylactic antiemetic therapy was repeated 12 h after therapy.

[^177^Lu]Lu-DOTA-MGS5 was administered intravenously via a dedicated peristaltic infusion pump system at a flow rate of 100 mL/h over a period of 15–20 min. The patient was discharged at the third day following therapy. The treatment was repeated approximately every four weeks.

#### Image acquisition protocol

Whole-body SPECT/CT after each therapy cycle was performed using a dual-head SPECT/CT system (NM/CT 870 DR, GE Healthcare, USA) approximately 24 h after injection of [^177^Lu]Lu-DOTA-MGS5. The acquisition consisted of 60 projections over 360° with an angular step of 6° and a dwell time of 20 seconds per projection. A medium-energy general-purpose collimator was used. The primary energy window was set to 187.2–228.8 keV and the scatter window to 169.1 - 186.9 keV. SPECT images were reconstructed using the manufacturer’s proprietary algorithms (OSEM with resolution recovery), using 2 iterations and 10 subsets, and no post-reconstruction filtering. The reconstructed matrix size was 128 × 128 with a pixel size of 4.42 mm. Photon scatter correction was performed using the dual energy window method, and attenuation correction was performed using a low-dose CT scan acquired at 120 kVp, 30 mAs effective, with CareDose4D activated. Whole-body planar imaging was performed in anterior and posterior views using the same system. The scan was conducted in supine position, feet-first orientation. The table traversed 1.14 m at a speed of 10 cm/min. The matrix size was 1024 × 256 with a pixel spacing of 2.40 mm.

For dosimetry, both planar and SPECT/CT imaging was performed at nominal time points of 30 min, 4, 24, 72, and 96 h post-injection (p.i.). The acquisition protocol for the first three time points followed the procedure described above. For the 72-and 96-hour scans, the dwell time per projection for SPECT acquisition was increased to 24 s, and the planar scan speed was reduced to 8 cm/min to compensate for the reduced count statistics at later time points.

#### Quantification and SPECT-SUV

The gamma camera sensitivity was calibrated according to the GE Healthcare-recommended protocol (DICOM Conformance Statement: NM General Purpose 600/800 Series, Rev. 15, 2018). A Petri dish containing a known activity of lutetium-177 was used as the calibration source, yielding a calibration factor of 5.2 cps/MBq. To correct for partial volume effects (PVE), recovery coefficients (RCs) were determined using an IEC-NEMA phantom with fillable spheres of varying diameters, and a Jaszczak phantom containing a 60 mm sphere (Data Spectrum Corporation, USA). For each sphere, RCs were calculated, and a logarithmic regression model was fitted to generate a recovery curve. For inter-cycle comparison of the SPECT-derived uptake, lesions were segmented on 24-hour p.i. SPECT images using a 41% isocontour threshold in Affinity 3.0 (Hermes Medical Solutions). Subsequently, SUV_max_ values normalized to body weight, were computed and corrected for PVE using the corresponding RCs.

## Results

### Fluorescence based receptor specific cell uptake

Cellular microscopy tracking of the fluorescent ligand in A431-CCK2R cells demonstrated rapid internalization of ATTO-488-MGS5. No uptake was visible in A431-mock cells imaged at 10 min and 60 min after incubation with ATTO-488-MGS5. Blocking experiments for the same time points showed that also pre-incubation with a 100-fold excess of DOTA-MGS5 prior to exposure to the fluorescent labeled ligand prevented the uptake of ATTO-488-MGS5, further confirming that the uptake occurs specifically through CCK2R (Figure 1B). Time-lapse fluorescence microscopy of the same field of view in A431-CCK2R cells revealed the dynamic internalization of ATTO-488-MGS5. Upon addition of the fluorescent ligand, rapid binding to the cell surface was observed. Within minutes, the fluorescently labeled ligand was internalized leading to progressive accumulation of ATTO-488-MGS5 in intracellular compartments over time, with significant intracellular fluorescent signal persisting even 4 h after incubation (Figure 1C).

### Short-and Long-Term Cytotoxic Effects of Radiation

The short term-induced irradiation effect was determined by analyzing the cell viability using the trypan blue exclusion method. The viability of A431-CCK2R cells irradiated with EBRT was dose-dependent, with the percentage of relative viability decreasing from 67.6 ± 5.2% at 2 Gy to 30.0 ± 3.2% at 8 Gy. The response of A431-CCK2R cells to treatment with [^177^Lu]Lu-DOTA-MGS5 (∼250 kBq, 2 nM/well) was comparable for 4 h and 24 h of incubation, with relative viability of 58.5 ± 3.3% for the 4 h and 55.5 ± 8.3% for the 24 h incubation period (Figure 2A). Morphological analysis showed that cell circularity remained consistent across all experimental conditions, indicating no major changes in overall cell shape. However, average cell diameter increased significantly in PRRT-treated cells compared to the control group. Also, in EBRT-treated cells the highest dose level of 8 Gy produced a significant increase in cell diameter.

**Figure 2.**
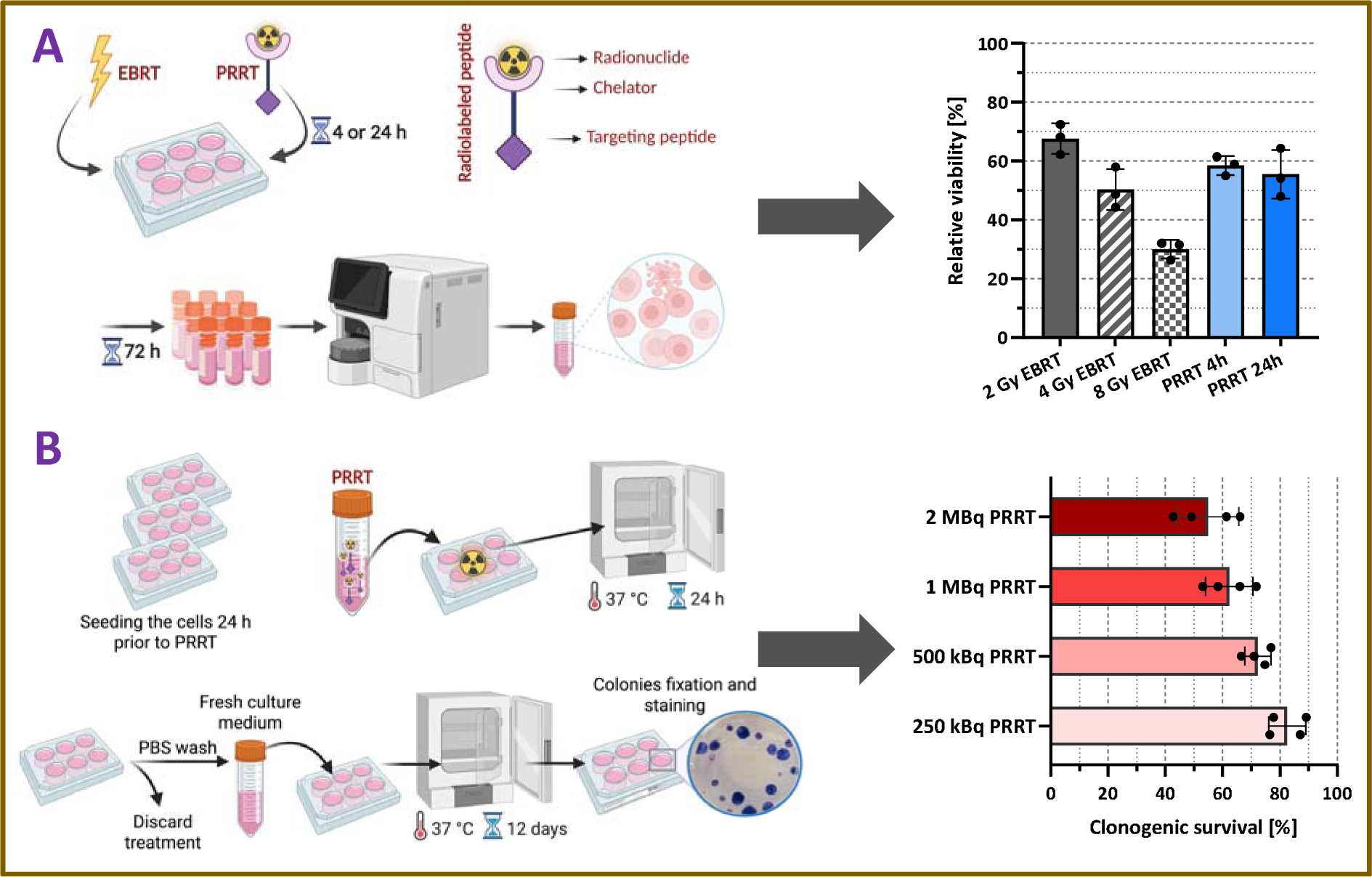
*In-vitro* radiation effect on A431-CCK2R cells. (A) Schematic illustration of the short-term cell viability assay using EBRT or PRRT and percentage of relative viability 72 h after exposure to different doses of EBRT and varying incubation periods with [^177^Lu]Lu-DOTA-MGS5 (n=3). (B) Schematic illustration of clonogenic assay and percentage of clonogenic survival 12 days after treatment with different doses of [^177^Lu]Lu-DOTA-MGS5 (n=4).

**Figure 3.**
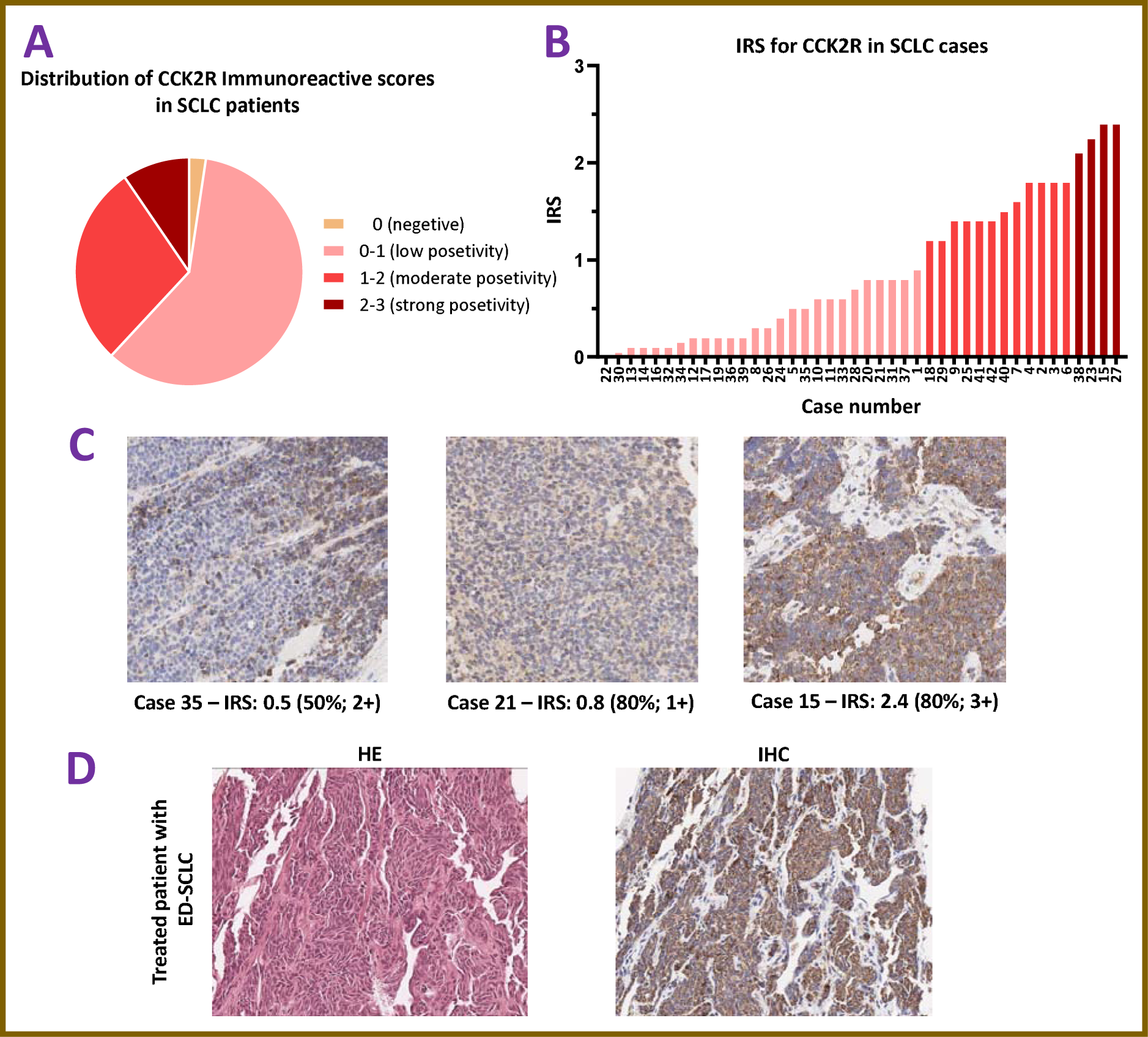
CCK2R immunoreactivity as analyzed on tissue specimens of patients with SCLC: (A) distribution of CCK2R immunoreactivity scores (IRS), (B) case by case visualization sorted by IRS, (C) exemplary images of IHC staining for scores 0.5 – 2.4. (D) Representative images of HE and IHC staining of patient biopsy samples from neoplasm of the left upper lobe of the lung of patient treated with [^177^Lu]Lu-DOTA-MGS5. Images are displayed at 400× magnification; brown staining indicating CCK2R expression.

In order to investigate the long-term effect of PRRT on A431-CCK2R cells, clonogenic assays were performed with different concentrations of [^177^Lu]Lu-DOTA-MGS5. Clonogenic survival of A431-CCK2R cells decreased in a dose-dependent manner following treatment with 250 kBq to 2 MBq [^177^Lu]Lu-DOTA-MGS5. The percentage of cell survival was reduced to 82.7 ± 6.5% at the dose of 250 kBq/well and decreased to 54.9 ± 10.9% at the dose of 2 MBq/well (Figure 2B).

### Immunohistochemistry

Immunohistochemical evaluation of tissue specimens from patients with SCLC revealed moderate to high CCK2R expression in more than one third of the specimens evaluated (38.10%, 16 patients out of 42). Immunoreactivity was mainly observed cytoplasmic with a granular staining pattern compatible with endosomal localization. Of the evaluated tissue specimens, one was negative with an IRS score of zero, 25 patients had an IRS score between 0 and 1, 12 patients had moderate CCK2R expression with an IRS score of 1 to 2, and 4 patients were strongly positive with an IRS score of 2 to 3 with the maximum IRS of 2.4 corresponding to 80% of tumor cells with strong positivity (Figure 3A-C).

Immunohistochemical evaluation was also performed for the first patient treated with [^177^Lu]Lu-DOTA-MGS5. As shown in Figure 3D, the biopsy sample from the bronchial mucosa of the left upper lobe of the lung from this patient was strongly positive for CCK2R expression with an IRS score of 2.1.

### Pharmaceutical formulation

A lower dose of [^177^Lu]Lu-DOTA-MGS5 with a radioactivity concentration of 105.2 MBq/mL and a final product activity of 1,505 MBq was synthesized for the patient-specific dosimetry study.

Subsequently, four different batches of [^177^Lu]Lu-DOTA-MGS5 with radioactivity concentration of 458.2 ± 10.0 MBq/mL and final product activity of 6507.3 ± 193.4 MBq were synthesized as therapeutic doses. Quality control of each individual batch confirmed radiochemical purity (RCP)>95% as analyzed by radio-HPLC. In addition, instant thin layer chromatography (iTLC) confirmed the presence of ≤ 1% free lutetium-177 and ≤ 2% colloidal lutetium-177. The quality control results for all therapeutic doses prepared for patient administration are shown in Supplementary Table 1. All batches produced complied with the defined specifications for the various parameters tested (appearance, pH, radioactivity concentration, radionuclide identity, identity, radiochemical purity, peptide content, apparent specific activity, ethanol content, bacterial endotoxins, and sterility) and met the required acceptance criteria for release.

### First patient experience of PRRT with [^177^Lu]Lu-DOTA-MGS5

Due to disease progression observed at the radiologic follow-up examination during fourth-line treatment with topotecan (Figure 4, a–c and d–f, respectively), and following confirmation of intense tracer uptake on [^68^Ga]Ga-DOTA-MGS5 PET/CT, the patient underwent a preliminary dosimetric evaluation with [^177^Lu]Lu-DOTA-MGS5 to calculate the absorbed dose to organs at risk and the tumor. After the intravenous administration of 1.5 GBq [^177^Lu]Lu-DOTA-MGS5, scintigraphic images were acquired at 30 min, 4 h, 24 h, 72 h and 96 h p.i. (Supplementary Figure 2A). As previously described, the patient-specific dosimetry evaluation revealed absorbed doses of 0.28 Gy/GBq to the kidneys, 0.022 Gy/GBq to the bone marrow, and 0.42 Gy/GBq to the stomach. Additionally, the mean absorbed dose to the tumor was 12.5 ± 10.2 Gy/GBq (range: 1.2–28 Gy/GBq) across five well-delineated lesions (25). The time–activity curves for kidneys, stomach wall, and two selected lesions were fitted and extrapolated using nonlinear regression. As shown in Supplementary Figure 2B, tracer clearance exhibited organ-and lesion-specific kinetics. The kidneys demonstrated a rapid initial decline in activity, consistent with fast clearance, while the CCK2R-positive stomach wall showed a slower and more prolonged washout. The thoracic wall lesion and gastrointestinal tract lesion displayed similar kinetic profiles, characterized by a gradual mono-exponential decrease in activity over time.

**Figure 4.**
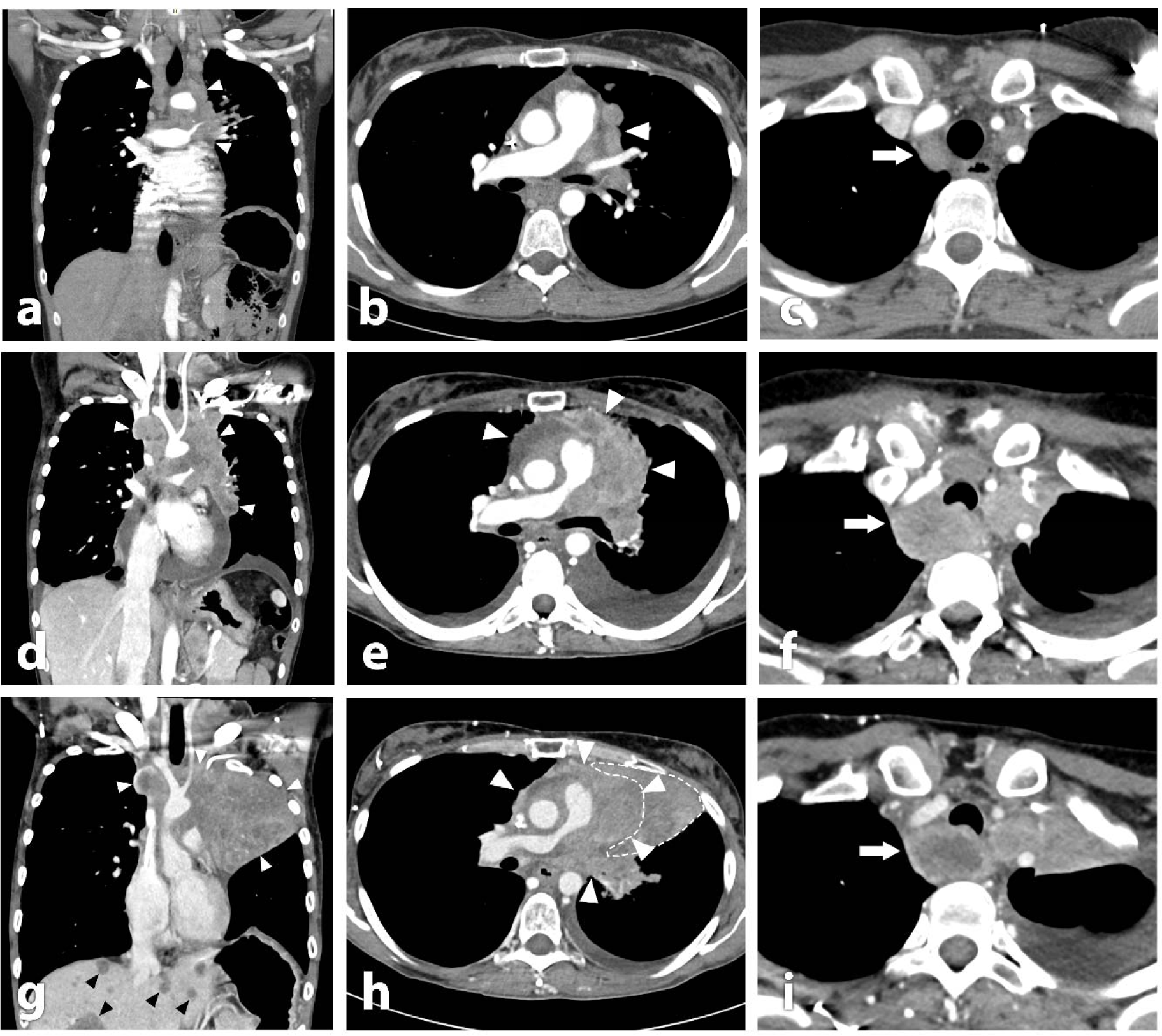
Coronal and axial computed tomography of the chest 4 months prior to (a-c), immediately prior to (d-f) and 3 months after treatment (g-i) with [^177^Lu]Lu-DOTA-MGS5. Over the time course progressive tumor extension (white arrowheads) into the mediastinum was observed, leading to increasing compression of mediastinal structures, illustrated by a progressive narrowing of the right pulmonary artery (b, e, h). Notice also areas of increasing tumor necrosis during growth, as demonstrated in a retrotracheal mass (white arrow; c, f, i) and progressive infiltration of the left main bronchus leading to paracardial atelectasis (dashed outline). At the last check-up, disseminated liver metastases was noticed (black arrowheads; g).

After confirming the treatment’s feasibility due to the low dose load to organs at risk, the patient proceeded with [^177^Lu]Lu-DOTA-MGS5 therapy. Prior to the start of treatment, the patient was alert, in good general condition, and reported no pain. Nausea was noted on an empty stomach. The patient’s weight was 66 kg with a BMI of 21.8 kg/m². Blood values, including white blood cell count, hemoglobin, and thrombocytes, as well as kidney parameters (creatinine 0.66 mg/dL, eGFR >60 mL/min/1.73 m²) and liver function tests (GOT 31 U/L, GPT 15 U/L, GGT 72 U/L), were all within the acceptable range to support PRRT. Tumor markers indicated a high neuron-specific enolase (NSE) level of 166.0 ng/mL, while chromogranin A was within the normal range.

About two years after initial diagnosis, the patient underwent four monthly cycles of [^177^Lu]Lu-DOTA-MGS5 therapy, with a cumulative administered activity of 17.2 GBq (mean activity per cycle: 4.3 GBq). The therapy was well-tolerated, with no acute or delayed adverse effects. Additionally, no hematological, renal, or hepatic toxicities were detected in laboratory tests performed during treatment period. NSE levels showed a slight reduction after the first three administrations but increased by the fourth administration, alongside a rise in chromogranin A levels (Supplementary Figure 3).

The patient’s performance, as measured by the Karnofsky Index, remained at 100% throughout treatment. Following the first administration, the patient experienced mild nausea. After the second administration, clinical symptoms showed improvement, although nausea and mild appetite loss persisted for a few days post-therapy, accompanied by a weight loss of approximately 2 kg. By the fourth administration, nausea and appetite improved further, although the patient reported a mild onset of dyspnea.

Serial coronal and axial fused SPECT/CT imaging acquired after each treatment cycle with [^177^Lu]Lu-DOTA-MGS5 demonstrated predominantly stable disease (Figure 6). The retrotracheal mass exhibited consistent uptake across all cycles, indicating stable metabolic activity. In contrast, the gastrointestinal tract lesion showed only a slight decline in SUV_max_, from 37 g/mL at baseline to 31 g/mL after the third cycle. The thoracic wall lesion exhibited a more marked decrease in SUV_max_, from 21 g/mL at dosimetry to 6 g/mL by the second cycle, with minimal change thereafter (Figure 7B). However, SPECT/CT imaging performed after the fourth cycle revealed new disseminated liver metastases, which were [^177^Lu]Lu-DOTA-MGS5-negative, indicating progressive disease (Supplementary Figure 4). Imaging after the last therapy cycle had to be performed on a different camera system and with a different protocol not allowing for a comparison of uptake values (for details see Supplementary Materials).

**Figure 6.**
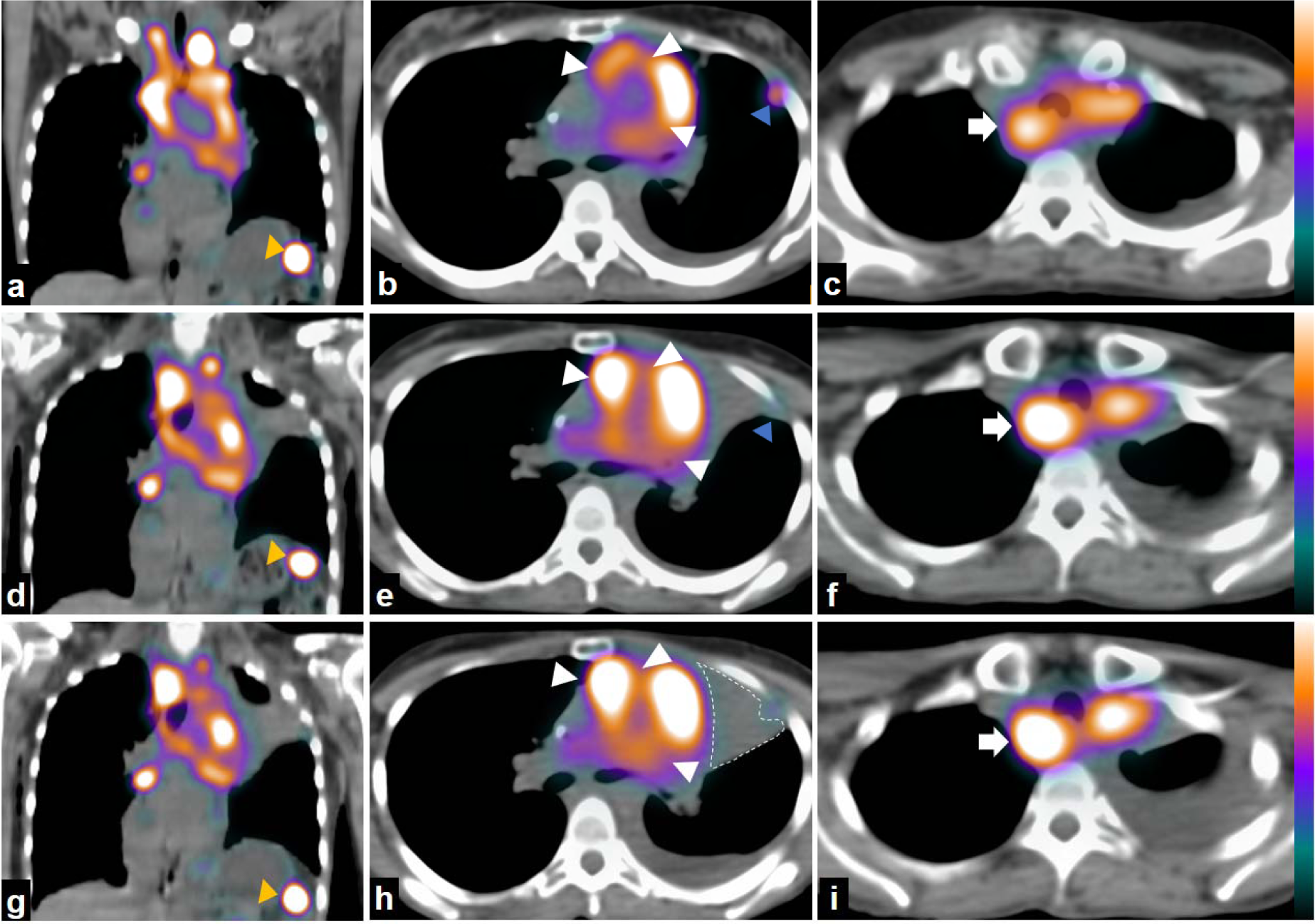
Coronal and axial fused single photon emission tomography/computed tomography (SPECT/CT) after the first (a-c), second (d-f) and third (g-i) cycle of treatment with [^177^Lu]Lu-DOTA-MGS5. Imaging showed substantial stable disease over time (white arrowheads) with a slight response in small tumor lesions such as those in the gastrointestinal tract (a, d, g - yellow arrowheads) and in the left thoracic wall (b, e, h - blue arrowheads)-the same ones used for dosimetry studies. The retrotracheal mass (white arrow; c, f, i) appears to demonstrate constant uptake during the course of treatment.

**Figure 7.**
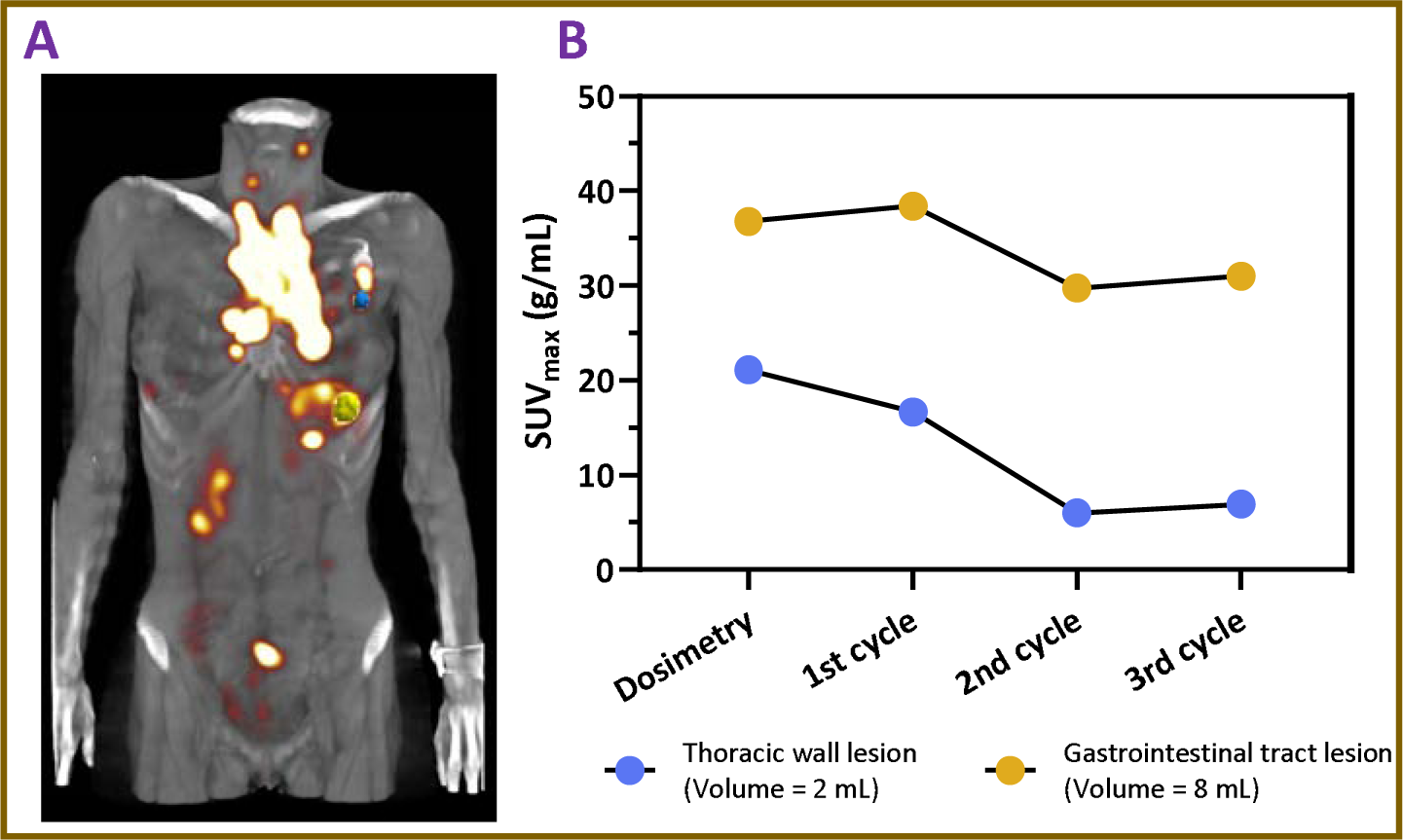
(A) Fused coronal view of a whole-body SPECT/CT scan acquired 24 h p.i. of [^177^Lu]Lu-DOTA-MGS5. The thoracic wall lesion is masked in blue, and the gastrointestinal tract lesion is masked in yellow. (B) A comparison of lesion uptake at 24 h p.i., indicated by SUV_max_, is shown for both well-delineated lesions during dosimetry and the first three treatment cycles.

The follow-up CT performed prior to the fourth cycle of PRRT demonstrated continued disease progression, with primary tumor progress leading to increasing compression of mediastinal structures, illustrated by a progressive narrowing of the right pulmonary artery, increasing central necrosis of the retrotracheal mass and progressive infiltration of the left main bronchus with paracardial atelectasis (Figure 4). New disseminated liver metastases were also noted together with further progression of lymph nodal metastasis (Figure 4, g-i).

PRRT was discontinued and fifth-line therapy with lurbinectedin was initiated. However, due to treatment-related toxicity and a marked decline in the patient’s general condition following the first cycle, the therapy was discontinued. With ongoing tumor progression, the patient passed away due to respiratory insufficiency and multiorgan failure one month later.

## Discussion

The scope of the present study was to provide further evidence on the applicability of [^177^Lu]Lu-DOTA-MGS5 for peptide receptor radionuclide therapy targeting the cholecystokinin-2 receptor. For this purpose specific preclinical testing was performed to further elucidate the receptor interaction and therapeutic efficacy. In dynamic fluorescent microscopy studies on A431-CCK2R cells incubated with Atto-488-MGS5, a rapid translocation from the cell surface to intracellular compartments leading to prolonged intracellular sequestration was confirmed, providing the basis for efficient receptor-specific tumor targeting. In the cell viability studies, the effect of 250 kBq of [^177^Lu]Lu-DOTA-MGS5 was laid between the effects of EBRT with 2 and 4 Gy. The observed significant increase in cell diameter found for PRRT-treated cells compared to control cells group suggests an early cytotoxic response to PRRT, consistent with known radiation-induced effects such as cellular stress, disruption of cell cycle progression, and swelling due to G2/M phase arrest. These morphological alterations may occur prior to measurable loss of cell viability and could reflect early signs of apoptosis or mitotic catastrophe (28). The performed clonogenic assays with increasing concentration of [^177^Lu]Lu-DOTA-MGS5 showed a dose-dependent effect on clonogenic survival suggesting that a higher level of irreparable DNA damage occurs at higher dose of [^177^Lu]Lu-DOTA-MGS5 which leads to long-term reproductive death. The dose-dependent decrease in clonogenic capacity is consistent with the mechanism of action of β-emitting radiopharmaceuticals, which induce lethal DNA damage through continuous low-dose irradiation, ultimately impairing the ability of cells to undergo multiple rounds of cell division (28, 29). To further elucidate the cellular effects of PRRT, we intend to conduct additional *in vitro* assays, such as the γH2AX assay, which will allow us to better characterize DNA double-strand break formation and repair dynamics, in addition to our current findings (30, 31).

To explore the applicability of this new personalized therapeutic approach in patients with SCLC, we have investigated the CCK2R receptor expression in tumor samples from patients with SCLC by immunohistochemistry. The analysis of 42 tumor specimens confirmed a moderate to high CCK2R expression in more than one third of the specimens evaluated supporting that [^68^Ga]Ga-DOTA-MGS5 can be used to target CCK2R in this patient group for selecting patients eligible for PRRT with [^177^Lu]Lu-DOTA-MGS5. As shown previously for somatostatin receptors, there is a moderate to strong correlation between somatostatin receptor expression and SUV_max_ of [^68^Ga]Ga-DOTA-TATE or [^68^Ga]Ga-DOTA-TOC PET scans in different tumor entities (9, 32, 33). No similar studies have been performed for CCK2R targeting so far. When investigating CCK2R expression in NETs by receptor autoradiography Reubi et al. reported a high incidence of CCK2R expression in MTC (92%, 22 of 24 samples), SCLC (57%, 8 of 14 samples), astrocytomas (65%, 11 of 17 samples) and stromal ovarian cancer (100%, 3 of 3 samples). In further studies, also insulinomas, vipomas, as well as some bronchial and ileal carcinoids, were suggested as potential CCK2R targets, especially in patients with low or no SSTR expression. In addition, also gastrointestinal stromal tumors (mean density of 8,641 dpm/mg) and leiomyosarcomas (mean density of 7,283 dpm/mg) express CCK2R at high density (34–36). Based on our immunohistochemical analysis in 38% of the SCLC tissue specimens analyzed, a high over-expression of CCKR could be confirmed, meeting the essential condition for the effectiveness of any receptor-based radionuclide therapy. In a new clinical trial which has recently been started in our center (EUCT n. 2024-514584-25-00) we will investigate the correlation of cholecystokinin receptor expression and uptake values of [^68^Ga]Ga-DOTA-MGS5 PET/CT imaging in patients with SCLC.

We could recently provide the first evidence of [^68^Ga]Ga-DOTA-MGS5-positive lesion uptake in a patient with ED-SCLC (23). The patient underwent dosimetric evaluation with [^177^Lu]Lu-DOTA-MGS5 in March 2024 (25), revealing high tumor dose in the range of 1.2–28 Gy/GBq combined with favorable pharmacokinetics leading to a low radiation burden of 0.28 Gy/GBq to the kidneys and 0.42 Gy/GBq to the stomach due to physiological uptake in this organ. Based on a tumor board decision a first therapeutic attempt with [^177^Lu]Lu-DOTA-MGS5 was undertaken in this patient. A treatment regimen with a dose of 4 GBq in intervals of one month was applied to this patient with extensive tumor progression. The patient was treated with a total of 4 cycles reaching a total administered activity of 17.2 GBq. This approach was chosen in the attempt to provide a palliative treatment to the patient, improving the patient’s quality of life while minimising potential side effects. Beside mild nausea and mild appetite loss for a few days post-therapy, the treatment was well tolerated by the patient, who reported a sustained state of psychological and physical well-being. Moreover, the patient maintained full autonomy in daily activities and showed subjective improvement in dyspnea and asthenia, mostly within the first three cycles of treatment. Of note, the patient received treatment with [^177^Lu]Lu-DOTA-MGS5 as a fifth-line therapy, following progression under various chemotherapy regimens. Therefore, the reduced occurrence of side effects is of particular importance. The post-therapy SPECT/CT scans after each treatment cycle demonstrated predominantly stable uptake suggesting partial stabilization of the disease. However, due to newly appearing disseminated liver metastases without [^177^Lu]Lu-DOTA-MGS5-uptake as well as further progression of the primary tumor PRRT was discontinued. Fifth-line therapy initiated thereafter was not tolerated by the patient and was also discontinued and the patient passed away in August 2024. However, with the use of the CCK2R-targeting theranostic pair, [^68^Ga]Ga-DOTA-MGS5 and [^177^Lu]Lu-DOTA-MGS5, we could offer an attractive and highly specific alternative treatment option to the patient, improving the patient’s quality of life. Such a theranostic approach combining imaging and therapy, allows for personalized treatment planning and monitoring (37). So far ^177^Lu-labeled peptides targeting CCK2R have been used for PRRT in patients with advanced MTC. In this patient group, the ^177^Lu-labeled CCK2R targeting agonist DOTA-(DGlu)_6_-Ala-Tyr-Gly-Trp-Nle-Asp-Phe-NH2 ([^177^Lu]Lu-PP-F11N) showed a favorable biodistribution with main accumulation in stomach and kidneys combined with a median absorbed dose for tumors of 0.88 Gy/GBq considered sufficient for a therapeutic approach (38). In a dose escalation study, 6 patients with advanced MTC were treated with up to 3-4 cycles of [^177^Lu]Lu-PP-F11N (3 x 6 GBq or 4 x 8 GBq) in intervals of 8-10 weeks without signs of dose limiting toxicity (38, 39). In another study, two patients with NETs showing sub-optimal uptake in SSTR targeting [^68^Ga]Ga-DOTA-TATE PET/CT imaging, were treated with the ^177^Lu-labeled CCK2R targeting agonist DOTA-(DGlu)_6_-Ala-Tyr-Gly-Trp-Met-Asp-Phe-NH2 ([^177^Lu]Lu-CP04), showing the potential theranostic application in patients with low SSTR-expressing NETs (40). The first patient treated with [^177^Lu]Lu-DOTA-MGS5 showed a tumor dose of 12.5 ± 10.2 Gy/GBq (range: 1.2–28 Gy/GBq), confirming the high potential of this new radiopharmaceutical in this patient group. The high tumor burden in the patient, consistent with a very advanced and progressed stage of the disease at the time of treatment, may have limited the long-term efficacy of the therapy. With increasing dedifferentiation NETs become more aggressive and do not express the receptor targets for therapy (41). This was possibly also the case in the patient treated with [^177^Lu]Lu-DOTA-MGS5. In future clinical trial (EUCT n. 2024-518039-12-00), we will investigate [^177^Lu]Lu-DOTA-MGS5 PRRT in larger patient cohort to identify patients who could benefit from this new treatment at an earlier stage of the disease.

## Conclusion

In this study, we demonstrated the receptor-mediated cellular internalization and strong intracellular retention of DOTA-MGS5 in CCK2R expressing cells. In addition, short-and long-term radiation effects of [^177^Lu]Lu-DOTA-MGS5 were demonstrated using different cell-based assays. The moderate to high CCK2R expression in SCLC supports the potential of a theranostic approach using DOTA-MGS5, with [^68^Ga]Ga-DOTA-MGS5 employed in PET/CT imaging for patient selection. This study provides the first proof of CCK2R-directed PRRT with [^177^Lu]Lu-DOTA-MGS5 in a patient with SCLC. The therapy was well tolerated, with no signs of acute toxicity leading to sustained state of well-being during the course of treatment. In further clinical studies, the therapeutic value and clinical applicability of this CCK2R-targeted approach needs to be verified in a larger patient cohort.

## Supplementary Materials

See additional file submitted.

## Data Availability

The datasets generated during and/or analysed during
the current study are available from the corresponding author on reasonable request.

## Acknowledgements

We gratefully acknowledge the guidance and training provided by Dr. Jean-Pierre Pouget and his team at the Institut de Recherche en Cancérologie de Montpellier in the implementation of the clonogenic assay. We would also like to thank the staff of the Department of Nuclear Medicine at the Medical University of Innsbruck for their assistance in performing the PRRT and SPECT/CT imaging studies.

## Author contributions

EG and TZ contributed to the study conception and design, and prepared the patient radiopharmaceutical doses. TZ performed the majority of the in vitro experimental work. JH contributed to the fluorescence microscopy studies. IS conceptualized and reviewed the cell toxicity studies. IP and KS performed the immunohistochemistry evaluations. AK performed the patient-specific dosimetry calculations. GDS, IV, GS, LG, DM, VM and JL were involved in patient management and interpretation of clinical results. TZ drafted the manuscript and prepared the figures. EG, LG, VM, IV reviewed and edited the manuscript. All authors read and approved the final version of the manuscript.

## Artificial Intelligence (AI) tools

This manuscript benefited from the assistance of the AI language model ChatGPT (OpenAI, https://www.openai.com/chatgpt) for language editing purposes. The final content was reviewed and edited by the authors to ensure accuracy and adherence to scientific standards.

