## Supplemental for "Peptide receptor radionuclide therapy targeting the cholecystokinin-2 receptor: Preclinical and first clinical experience in small cell lung cancer"

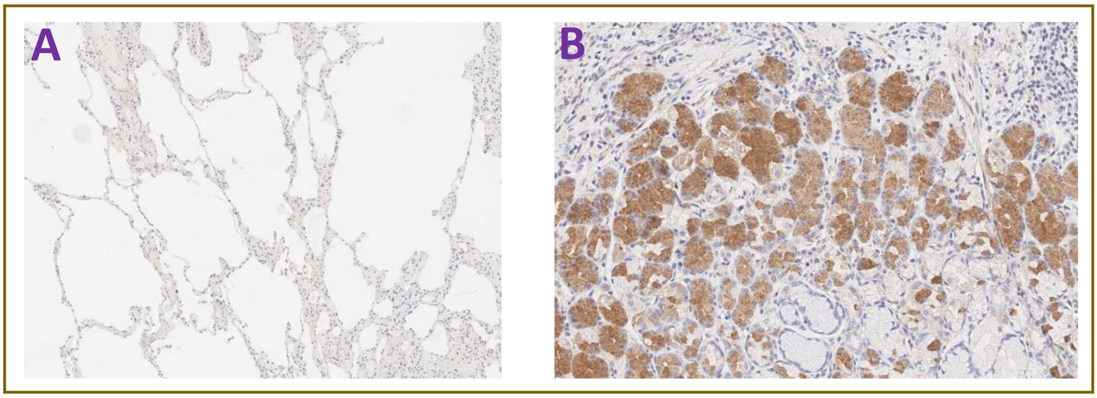


**Supplementary Figure 1.** CCK2R immunoreactivity; (A) normal lung tissue lacking CCK2R expression (magnification: ×100) (B) human stomach tissue, known to physiologically express CCK2R (magnification: ×300).

**Supplementary Table 1.** Acceptance criteria and quality control results of four individual therapeutic batches of [$^{177}$Lu]Lu-DOTA-MGS5

| Parameter | Method | Limits | Mean ± sd |
|---|---|---|---|
| Appearance | Visual inspection | Clear, colorless solution with no visible particulates | Conforms |
| pH | Indicator strip | 5-7 | 6 |
| Volume | Graduated vial (mL) | 10-20 | 14.2 ± 0.2 |
| Activity of the final product | MBq | >1000 | 6507.3 ± 193.4 |
| Radioactivity concentration | MBq/mL | <700 | 458.2 ± 10.03 |
| Radionuclide identity | Gamma-ray spectrometry (113 and 208 kev) | conforms | conforms |
| Identity of [$^{177}$Lu]Lu-DOTA-MGS5 | HPLC (comparison with reference $^{nat}$Lu-DOTA-MGS5) | 0.9-1.1 | conforms |
| Radiochemical purity | RCP (HPLC) | ≥95% | 97.9 ± 0.2 |
| Free lutetium-177 | TLC (0.1 sodium citrate pH 5) Rf 0.8-1.0 | <1% | 0.12 ± 0.08 |
| Radiocolloid | TLC (1 M ammonium acetate/methanol; 1/1); Rf 0-0.3 | <2% | 0.09 ± 0.05 |
| Limit test for peptide content | HPLC (UV) | ≤100 | 69.14 ± 33.01 |
| Apparent specific activity | MBq/µg | >20 | 65.07 ± 1.93 |
| Ethanol content | Gas chromatography (v/v) | ≤10% | 6.05 ± 3.23 |
| Bacterial endotoxins | LAL test (EU/V) | <175 | <37 |
| Sterility | Ph. Eur. | sterile | Sterile |
| Injected patient dose | MBq | - | 4321.3 ± 213.2 |
| Injected patient volume | Graduated vial | - | 9.4 ± 0.6 |

### Time-Activity Curve Fitting

Extrapolated time-activity curves were generated by fitting activity data obtained from quantitative SPECT imaging, normalized to the injected activity. A tri-exponential function was used for the kidneys, while a bi-exponential model (with $k_3 = 0$) was applied for the stomach wall and the two lesions.

Subsequently, the fitted curves were scaled to percent injected activity and normalized to organ or lesion mass (in grams), resulting in the following expression for the relative activity:

$$\boldsymbol{Activity}\left(\%\frac{IA}{g}\right) = \left(k_1 \times e^{-\lambda_1 t} + k_2 \times e^{-\lambda_2 t} + k_3 \times e^{-\lambda_3 t}\right) \times \frac{100}{mass(g)}$$

Fitting was performed using nonlinear regression in Microsoft Excel, using the Solver add-in based on a least-squares approximation. This model was used to describe the kinetics of tracer clearance and to extrapolate time-activity data beyond the final measurement time point.

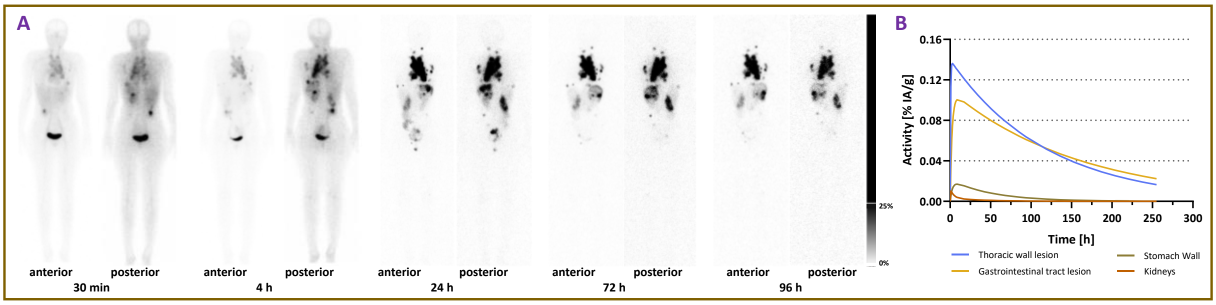


**Supplementary Figure 2.** (A) Representative serial whole-body planar images of a patient with ED-SCLC following intravenous administration of 1.5 GBq [$^{177}$Lu]Lu-DOTA-MGS5. (B) Time–activity fitted curves extrapolated for two lesions, as well as for the stomach and kidney, were derived from the serial scans.

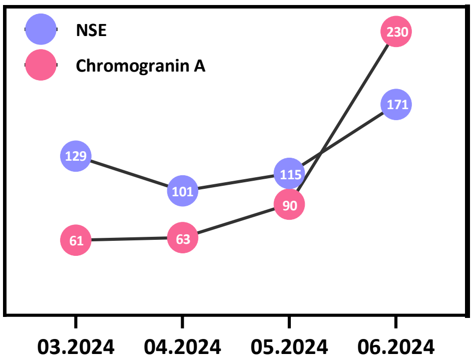


**Supplementary Figure 3.** Trend of tumor markers during PRRT with [$^{177}$Lu]Lu-DOTA-MGS5 (reported as ng/mL).

#### Image acquisition protocol after the last therapy cycle

Whole-body SPECT/CT after the last therapy cycle was performed using a dual-head SPECT/CT system (Symbia T, Siemens) 3.5 hours post-injection. The acquisition consisted of 64 projections over 360° with an angular step of 5.625° and a dwell time of 20 seconds per projection, using a medium-energy low-penetration (ME) collimator. Energy windows were set at 15% width around the photopeaks at 113 and 208 keV, each supplemented with upper and lower scatter windows for scatter correction. The SPECT images were reconstructed using the manufacturer's proprietary algorithm Flash 3D (OSEM with resolution recovery), applying 8 iterations and 4 subsets, and Gaussian filtering (FWHM = 9 mm). The reconstructed matrix size was 128 × 128 with a pixel size of 4.80 mm. Attenuation correction was performed using a low-dose CT scan acquired with a tube voltage of 130 kVp and an effective exposure of 17 mAs. Whole-body planar imaging was performed in anterior and posterior views using the same system. The scan was conducted in supine position, feet-first orientation. The table traversed 2 m at a speed of 15 cm/min. The matrix size was 1024 × 256 with a pixel spacing of 2.40 mm.

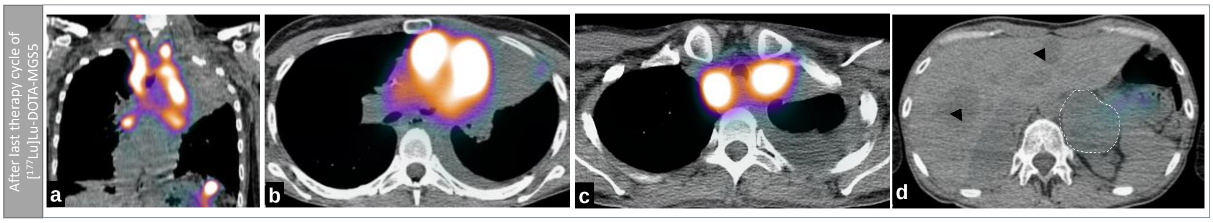


**Supplementary Figure 4.** Coronal and axial fused single photon emission tomography/computed tomography (SPECT/CT) after the fourth cycle of treatment with [$^{177}$Lu]Lu-DOTA-MGS5. Next to stable uptake in CCK2R-positive disease new disseminated liver metastases without [$^{177}$Lu]Lu-DOTA-MGS5 uptake was noticed in the CT scan (black arrowheads; d).
